# Evaluating the effect of COVID-19 on dispensing patterns: a national cohort analysis

**DOI:** 10.1101/2021.02.15.21251552

**Authors:** Fatemeh Torabi, Ashley Akbari, Laura North, Daniel Harris, Gareth Davies, Mike Gravenor, Rowena Griffiths, Jane Lyons, Neil Jenkins, Andrew Morris, Julian Halcox, Ronan A. Lyons

**Affiliations:** Population Data Science and Health Data Research UK, Swansea University; Swansea University; NHS Wales Shared Services Partnership

## Abstract

**Background:** Medication prescribing and dispensing often regarded as one of the most effective ways to manage and improve population health. Prescribed and dispensed medications can be monitored through data linkage for each patient. We hypothesised that changes in patient care resulting from COVID-19, changed the way patients access their prescribed medication.

**Objective:** To develop an efficient approach for evaluation of the impact of COVID-19 on drug dispensing patterns.

**Methods:** Retrospective observational study using national patient-level dispensing records in Wales-UK. Total dispensed drug items between 01-Jan-2016 and 31-Dec-2019 (counterfactual pre-COVID-19) were compared to 2020 (COVID-19 year). We compared trends of dispensed items in three main British National Formulary (BNF) sections(Cardiovascular system, Central Nervous System, Immunological & Vaccine) using European Age-Standardized rates. We developed an online tool to enable monitoring of changes in dispensing as the pandemic evolves.

**Result:** Amongst all BNF chapters, 52,357,639 items were dispensed in 2020 compared to 49,747,141 items in 2019 demonstrating a relative increase of 5.25% in 2020(95%CI[5.21,5.29]). Comparison of monthly patterns of 2020 and 2019 dispensed items showed a notable difference between the total number of dispensed drug items each month, with an average difference (D) of +290,055 and average Relative Change (RC) of +5.52%. The greatest RC was observed in a substantial March-2020 increase (D=+1,501,242 and RC=+28%), followed by second peak in June (D=+565,004, RC=+10.97%). May was characterised by lower dispensing (D=-399,244, RC=-5.9%). Cardiovascular categories were characterised, across all age groups, by dramatic March-2020 increases, at the epidemic peak, followed by months of lower than expected dispensing, and gradual recovery by September. The Central Nervous System category was similar, but with only a short decline in May, and quicker recovery. A stand-out grouping was Immunological and Vaccine, which dropped to very low levels across all age groups, and all months (including the March dispensing peak).

**Conclusions:** Aberration in clinical service delivery during COVID-19 led to substantial changes in community pharmacy drug dispensing. This change may contribute to a long-term burden of COVID-19, raising the importance of a comprehensive and timely monitoring of changes for evaluation of the potential impact on clinical care and outcomes

## Introduction

The novel coronavirus disease 2019 (COVID-19) pandemic has resulted in unprecedented changes on health care services provision (1). While technology has provided an increase in telephone or virtual appointments there has been an overall net reduction in primary care appointments (2). Increase in demand for essential medicines coupled with complex medicines supply chain issues, enforcement of social distancing, quarantine, and self-isolation has impacted the prescribing and dispensing of medicines (3)(4).

Studies using survey and patient data, reported an observed change in medication use during the pandemic (5)(6). Changes in service delivery have been monitored through multiple national audits which often lag considerably behind in real-time (7)(8). There is an urgent need to focus appropriate resources on optimisation of medication dispensing monitoring systems, especially in vulnerable patient groups; for example those receiving immunosuppressive drugs (9).

Electronic dispensing records holding information on all dispensed prescriptions in primary care, provide a unique opportunity to monitor dispensing trends. We aimed: 1) to measure the general impact of COVID-19 on dispensing patterns; 2) create a national research ready dataset of all primary care dispensing records for the whole population of Wales and 3) to design and implement a tool for monitoring and visualisation real-time trends.

## Method

### Study design and data sources

We conducted a retrospective observational study, accessing national patient-level primary care dispensing data using the SAIL Databank (10)(11). SAIL holds the Welsh Dispensing DataSet (WDDS), which records information on all national health service (NHS) prescription items dispensed by primary care dispensing contractors (community pharmacies, dispensing general practices, general practices that personally administer prescription medication and dispensing appliance contractors) from 2016, available on a monthly basis currently to end of September 2020. (12)

The WDDS data is provided to the SAIL Databank by NHS Wales Shared Services Partnership (NWSSP). The data is captured from prescriptions submitted to NWSSP, on a monthly basis, by all primary care dispensing contractors to claim remuneration for dispensing in accordance with the provisions of the National Health Service (Pharmaceutical and Local Pharmaceutical Services) Regulations 2013. The WDDS dataset is limited to NHS prescriptions containing a 2D matrix barcode i.e. those that are produced by clinical systems in GP practices in Wales. It therefore covers 94% of prescriptions dispensed and subsequently submitted for reimbursement. Medications for Welsh residents that are dispensed against non-barcoded prescriptions or dispensed outside Wales are not included in this dataset. These data are made available to SAIL on a monthly basis with approximately a 6-week lag to allow for official reporting and quality assurance processes (13).

Dispensing data from 1^st^ of January 2016 to 31^st^ of December 2019 (counterfactual pre-COVID-19, “C16 cohort”) were compared to data from 1st January 2020 to the 31^st^ of September 2020 (COVID-19 year, “C20 cohort”). For all people who were alive and resident in Wales with a record in the two purpose-built residency spine datasets (13), drug dispensing history was retrieved from patient-level linked dispensing record using anonymized linkage fields, and the date of dispensing of the drugs.

Dispensed items in WDDS are originally coded in Dictionary of Medicines and Devices (DM+D), which we mapped at a per code level to their corresponding British National Formulary (BNF) code(s) using an extract of mappings from National Health Service Business Services Authority (NHS-BSA) services published on December 2020 (14). Where the BSA mapping is not defined, NHS Terminology Reference-Data Update Distribution (known as TRUD) tables were used to map DM+D codes to 7^th^ character BNF codes (15) (see Supplementary 1 for more details on mapping).

Data were aggregated by month and year, for items in each British National Formulary (BNF) chapter. We compared dispensing rates between the C16 and C20 cohorts for the three main BNF chapters: I) Cardiovascular Systems, II) Central Nervous System and III) Immunological Products & Vaccines. We calculated age-standardised dispensing rates per 100,000 population. Five-year age bands were used with upper bound of all those aging 90 years or more using the 2013 European Standard Population (ESP) (16). Dispensing trends were assessed based on total quantity of dispensed items for each BNF chapter (17) per year and the number of patients who have had at least one item dispensed, as documented in their WDDS records.

### Statistical analysis

Per person dispensing rates were calculated based on the total number of dispensed items and total number of patients with a dispensing record on each period. Dispensing rates were calculated for 2020 and the counterfactual period of 2016 to 2019. The difference (D) and relative changes (RC) were calculated for comparison of monthly trends in dispensing between 2020 records and 2019. All statistical procedures are performed in R (v3.5) and the codes are available at https://github.com/SwanseaUniversityMedical/WDDS.

### Interactive tool

We developed an interactive tool using R shiny (18) which allows dynamic monitoring of trends in the current year and enables comparison between the most up to date data with previous years’ patterns. The dashboard is accessible from https://wdds.ml/.

## Results

### Cohort curation

3,139,662 patients with a total of 353,516,236 dispensing records entered in the WDDS were linked in two COVID-19 e-cohorts (13). This comprised 87.7% of Welsh residents between 2016 and 2019 (C16) who had at least one dispensing record in WDDS and could be successfully linked. The percentage for those with residency record at the start of 2020 (C20) was 93.5%. A mapping rate of 99.7% was achieved for mapping all dispensed records from DM+D to BNF codes for individuals in both the C16 and C20 cohorts who had linked data available for analysis (Figure 1 & Supplementary 1).

**Figure 1:**
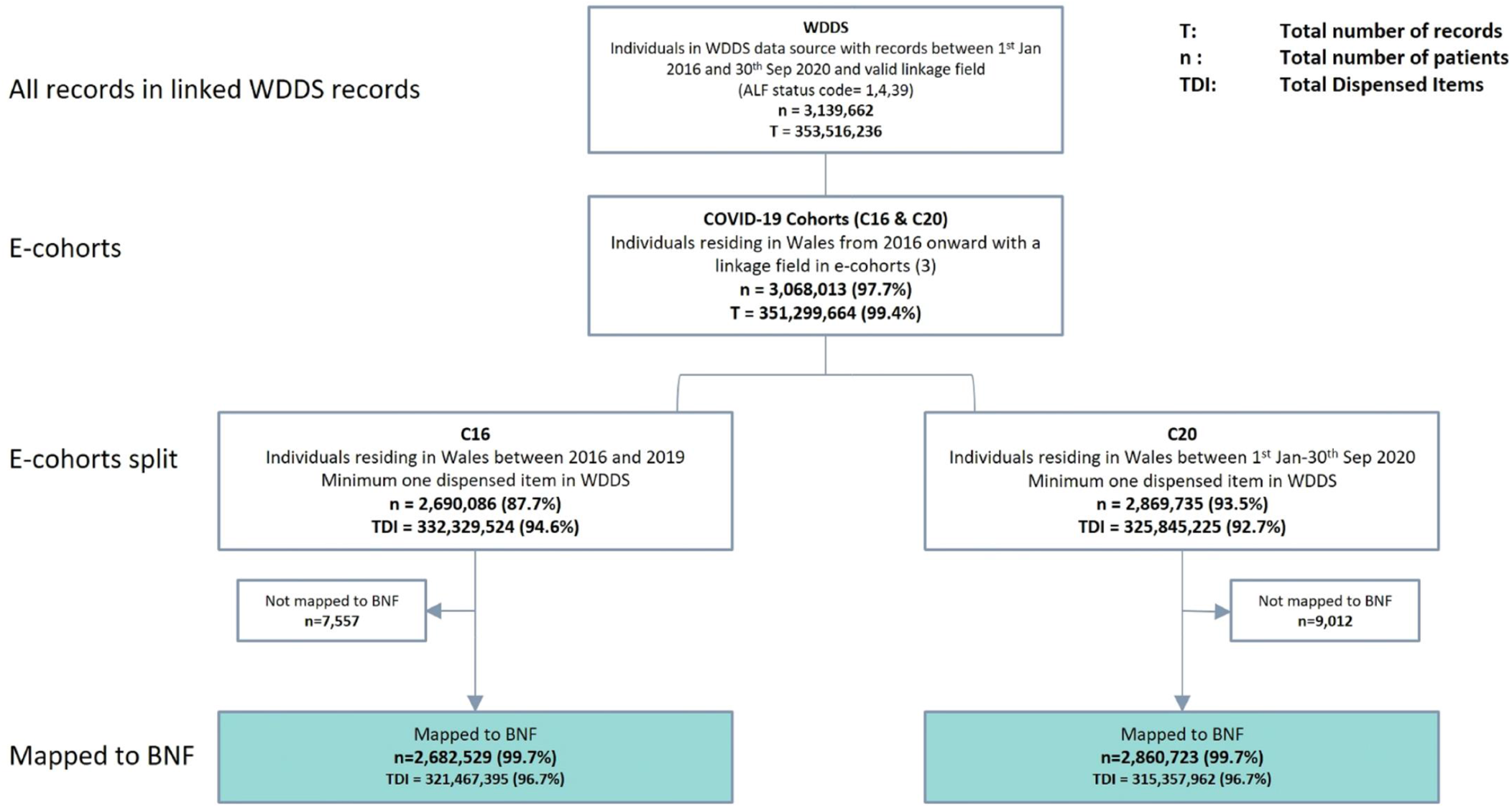
CONSORT of cohort extraction and number of matched mapped DM+D codes to BNF

### Drug dispensing trends

There was a 5.25% increase in total dispensed items in 2020 compared to the previous year (52,357,639 vs 49,747,141; 95% CI [5.21, 5.29]); while, overall rate of dispensed items per person remained unchanged: 26.25 items were dispensed per individual in 2019 and 25.92 in 2020 representing an Difference (D) of −0.3 and a Relative Change (RC) of −0.01% (Table 1.a).

**Table 1-a:** number of dispensed items per year

| Year | Number of dispensed items<br>(up to end of Sep) | Number of patients with a dispensed record<br>(up to end of Sep) | Dispensed Rate per person | Number of dispensed items<br>(Full year*) | Number of patients with a dispensed record<br>(Full year) | Dispensed Rate per person |
| --- | --- | --- | --- | --- | --- | --- |
| **2020** | **52,357,639** | **2,020,298** | **25.92** | **69,810,185** | **2,057,953** | **33.92** |
| 2019 | 49,747,141 | 1,895,365 | 26.25 | 66,539,725 | 1,998,607 | 33.29 |
| 2018 | 48,798,853 | 1,940,080 | 25.15 | 65,591,106 | 2,042,733 | 32.11 |
| 2017 | 46,554,918 | 1,978,408 | 23.53 | 62,522,297 | 2,090,082 | 29.91 |
| 2016 | 45,501,194 | 2,045,059 | 22.25 | 60,739,546 | 2,155,068 | 28.18 |

\*Presented numbers include estimate counts for 3 remaining months(Oct, Nov and Dec) of 2020

Comparison of monthly patterns of dispensed items between 2020 and 2019 showed an a notable difference between the total number of dispensed drug items each month, with an average difference of 290,055 and average RC of 5.52%. In the first two months of 2020, total number of dispensed items were 6% higher than 2019 (average D = +318,939, average RC =+6%). In March 2020, the total number of dispensed items was 28% higher than 2019 (D=+1,501,242, RC= +28.05%). Fewer items were dispensed in April 2020 than April 2019 (D= − 23,767, RC= −0.42%) and a further negative fall was observed in May 2020, where the total number of dispensed items was 5.88% lower than in May 2019 (D= −339,244, RC= −5.88). A second peak in total number of monthly dispensed items was observed in June of 2020 (D=+565,004, RC= +10.97%). Although the number of dispensed items increased in June, this was followed by a further reduction of total numbers in subsequent months with an average of 2.69% lower dispensing in July and August of 2020 compared to the same periods in 2019 (average D= −153,795, average RC= −2.69). A third peak representing a 9.7% greater number of dispensed items was observed in September 2020 compared to September 2019 (D=+529,442, RC= +9.75%) (Table 1.b).

**Table 1.b:**
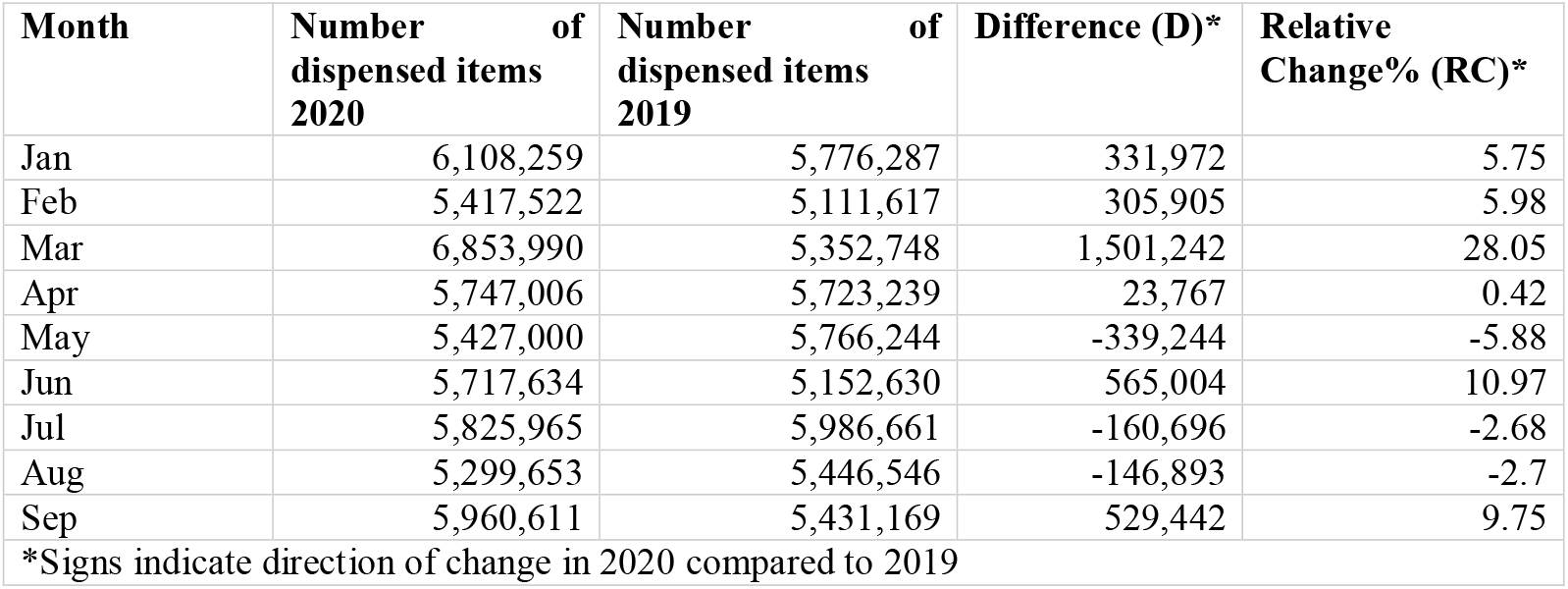
number of dispensed items per month of the year

| Month | Number of dispensed items<br>2020 | Number of dispensed items<br>2019 | Difference (D)* | Relative Change% (RC)* |
| --- | --- | --- | --- | --- |
| Jan | 6,108,259 | 5,776,287 | 331,972 | 5.75 |
| Feb | 5,417,522 | 5,111,617 | 305,905 | 5.98 |
| Mar | 6,853,990 | 5,352,748 | 1,501,242 | 28.05 |
| Apr | 5,747,006 | 5,723,239 | 23,767 | 0.42 |
| May | 5,427,000 | 5,766,244 | -339,244 | -5.88 |
| Jun | 5,717,634 | 5,152,630 | 565,004 | 10.97 |
| Jul | 5,825,965 | 5,986,661 | -160,696 | -2.68 |
| Aug | 5,299,653 | 5,446,546 | -146,893 | -2.7 |
| Sep | 5,960,611 | 5,431,169 | 529,442 | 9.75 |

\*Signs indicate direction of change in 2020 compared to 2019

Overall, comparison of relative changes in dispensing for 2020 vs 2019, between the first peak in dispensing in March (RC=+28.05%), the second peak in June (RC=+10.97%) and third peak in September (RC=+9.75%) indicates a progressive return towards pre-COVID-19 monthly dispensing patterns.

The observed increase in in March was consistent for items in the majority of BNF chapters, except for items in the *Anaesthesia* and the *Immunological products & Vaccines* and *Dressings* chapters, where the dispensing patterns were different to the overall patterns (Figure 2. For dynamic comparisons of all years, please see Supplementary Figures 2.1 & 2.2 or visit out online dashboard at http://wdds.ml/).

**Figure 2.**
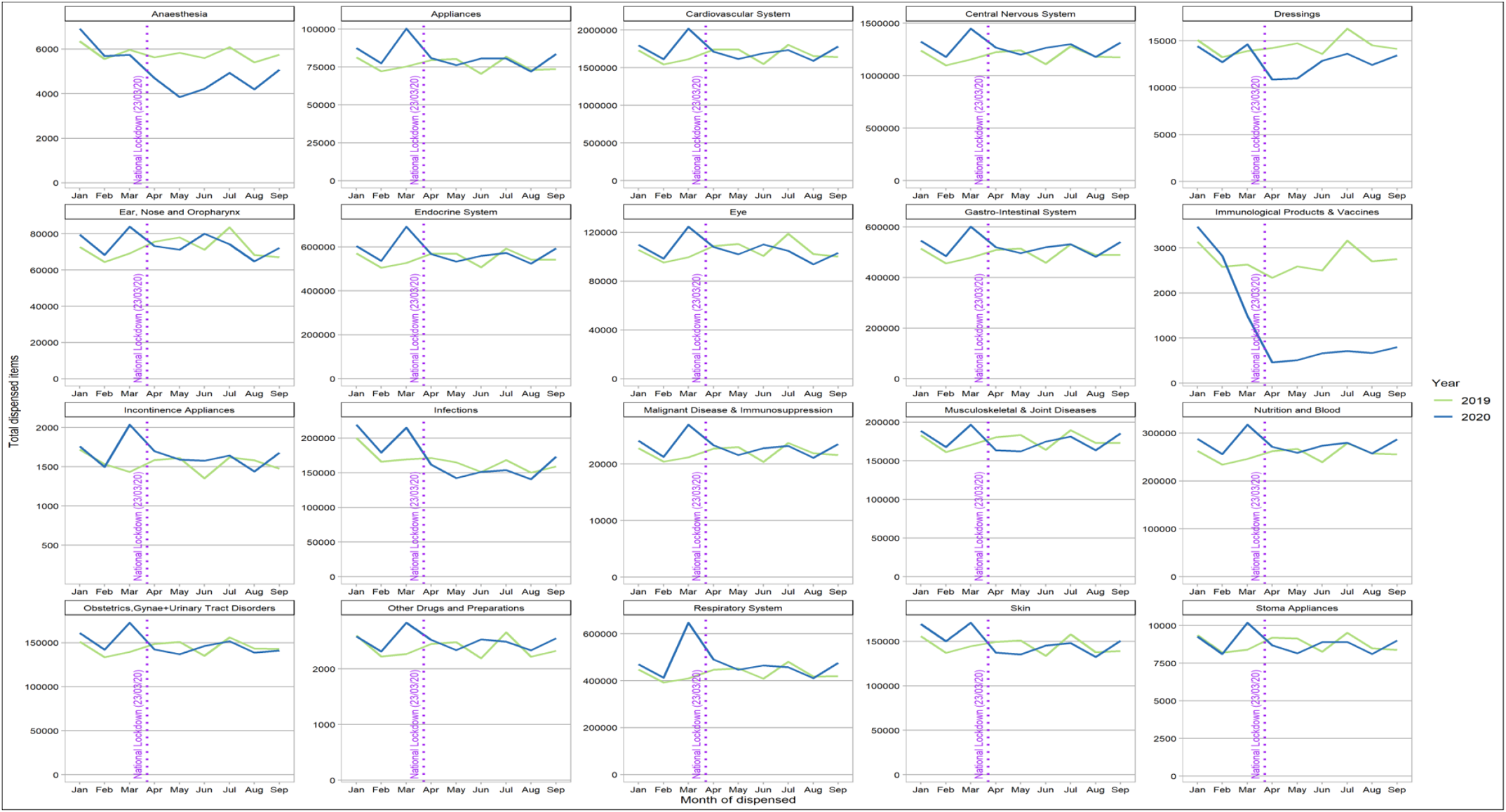
Number of dispensed items per BNF chapter per month – **in 2020 vs 2019** (numbers of each chapter are individually scaled)

### Trends of drug dispensing rates in three selected major BNF sections over years

Using age standardised dispensing rates, we focused on the comparison between 2019 and 2020 for Cardiovascular Systems (CVS), Central Nervous Systems (CNS) and Immunological Products and Vaccines (IPV). All three groups showed similar trends between the years for January and February. For CVS, dispensing increased considerably during March, however for subsequent months there was generally reduced dispensing, until recovery to 2019 levels in September (Figure 3 – Supplementary Figure 3.1). For CNS, the March 2020 peak was again observed, however only May appeared to show significant reductions compared to 2019 (Figure 4 – Supplementary Figure 3.2). The notable outlier was IPV. Although the 2020 dispensing patterns for January and February mirrored 2019 level, for all other months (and across all age groups) there was a large decline (Figure 5 – Supplementary Figure 3.3).

**Figure 3.**
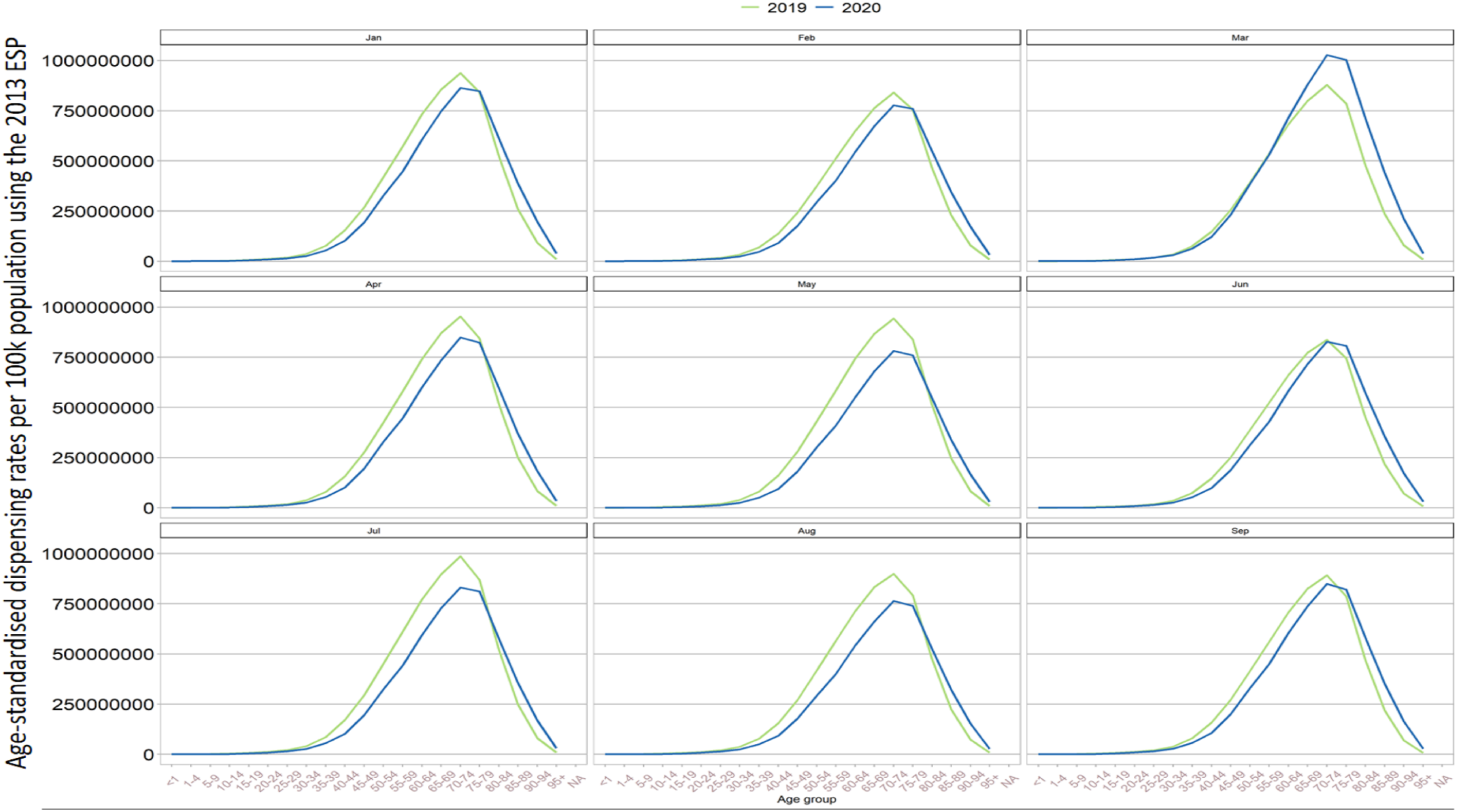
Age-standardized dispensing rates for **<u>cardiovascular system</u>** in per 100,000 pop’n per year and month for 2020 vs 2019

**Figure 4.**
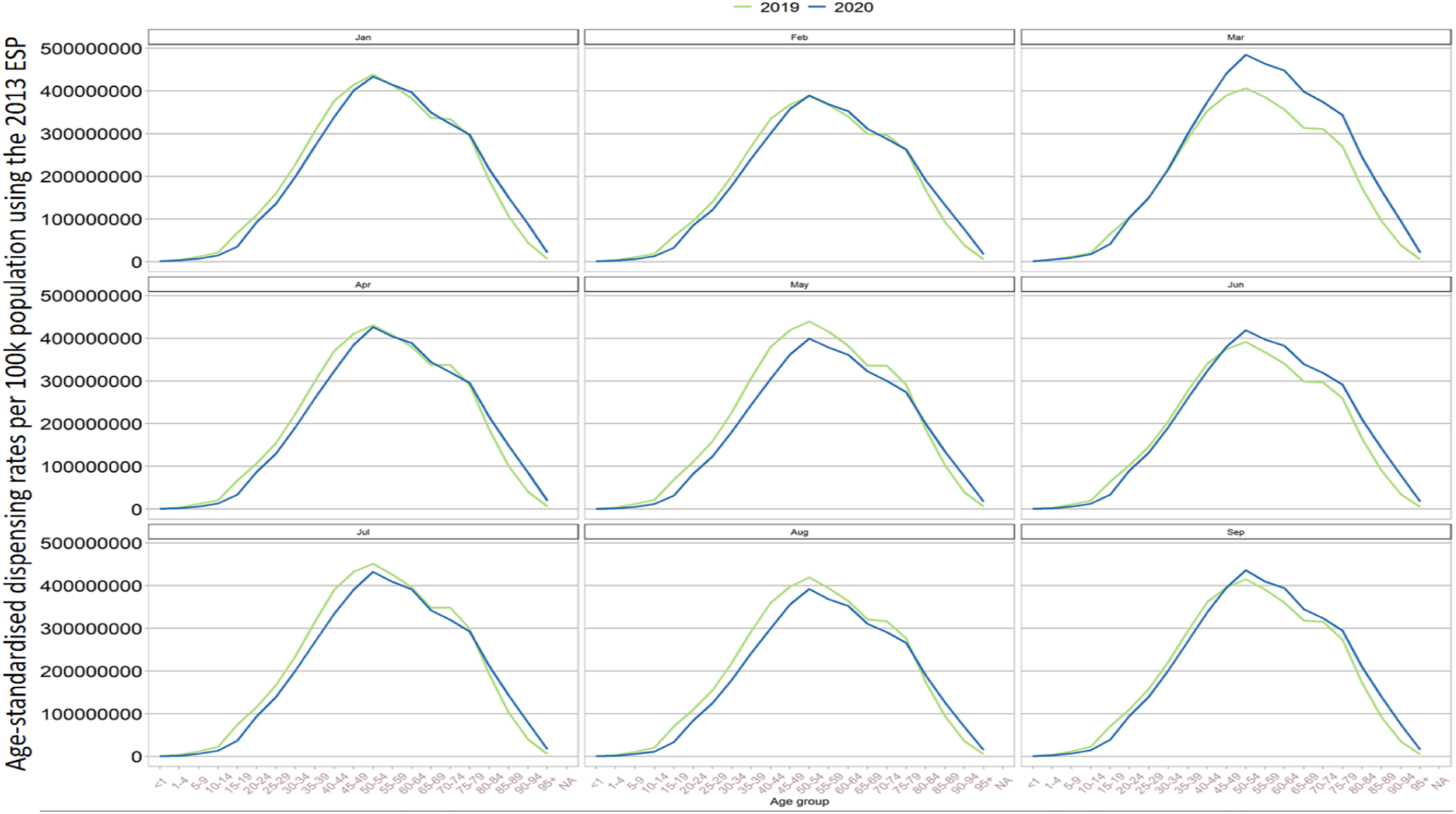
Age-standardized dispensing rates for **<u>Central Nervous System</u>** in per 100,000 pop’n per year and month for 2020 vs 2019

**Figure 5.**
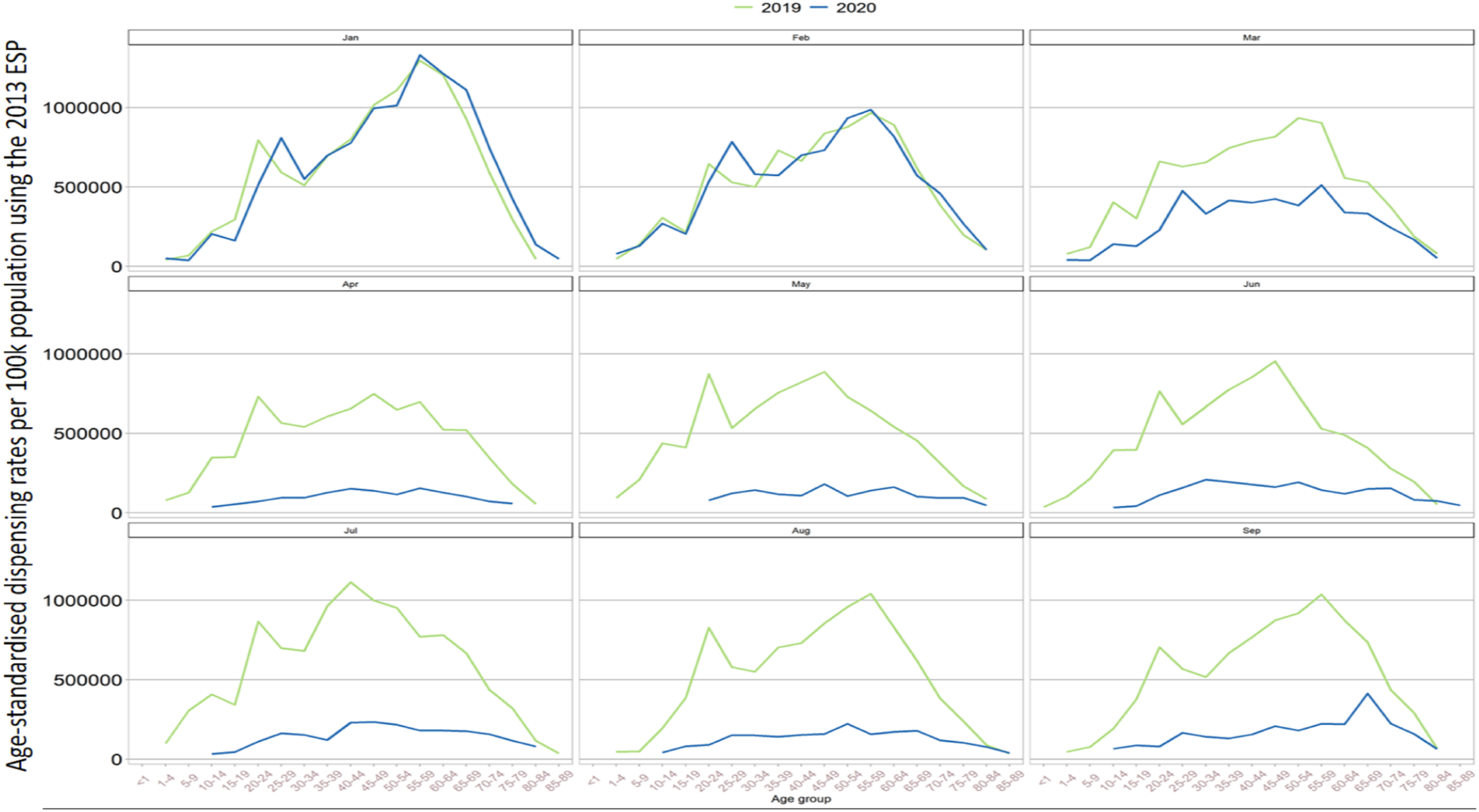
Age-standardized dispensing rates for **<u>Immunological Products & Vaccines</u>** in per 100,000 pop’n per year and month for 2020 vs 2019

## Discussion

### Summary of findings

This rapid assimilation of national dispensing data in Wales resulted in construction and curation of a research ready linked dataset. Our data have shown in detail, over all BNF chapters, changes in patterns of drugs dispensed during COVID-19 period. In general, the peak in total numbers of dispensed items coincided with the first UK national lockdown, at the first peak of the epidemic in March 2020. Our web application tool was developed for this comparative analysis using aggregated level data to enable timely monitoring of dispensing trends over time as the care system adapts to the evolving challenges of the pandemic.

We observed a dramatic reduction in the dispensing of Immunological Products & Vaccines at the start of lockdown in COVID-19 period. While routinely dispensed vaccines in pharmacies are mainly related to travel - such as yellow fever and rabies, the demand for these have naturally been reduced during the COVID-19 period. Similarly, obtaining vaccines from pharmacies through practice nurses prior to house visits may have been less frequent during COVID-19 lockdown periods. The current COVID-19 pandemic has perturbed routine immunisation and has led to dramatic shifts in the dispensing of Immunological Products & Vaccines (19)(20).

While our observation matches to what has been reported in other studies such as Kaye et al. (5); the increase of dispensed items in March cannot solely be considered as a better adherence. Our data showed that this peak occurred in most drug categories and was followed by a dip in the following two months (lockdown period) which is suggestive of batch-dispensing in preparation of the national lockdowns. It is also likely that patients may have had spare prescriptions to hand and felt an urgent to get them dispensed.

### Strengths and limitations

The patient level data used in our study are available as a monthly extract to SAIL Databank at the patient level; this provides the opportunity to use aggregated level data for monitoring trends in near real-time, as well as exploring the effect of patient-level factors such as age, sex, socioeconomic status and ethnicity. This mechanism provides the powerful opportunity to using these data not only to monitor and evaluate prescribing and dispensing trends in near real-time, but also to explore the effect of changing treatment patterns on outcomes considering a comprehensive selection of patient-level factors. Our study developed a version of the linked dispensing data which was has mapped DM+D codes to the BNF codes and chapters. This provides an opportunity to further investigate trends within each drug category, as well as wider collaboration across harmonised data sources.

## Conclusion

Dispensing patterns can be used as a proxy measure for monitoring community and population level effects of COVID-19 on healthcare services. By providing a unique and transparent system for continuous monitoring of drug prescribing and dispensing, this approach offers an unprecedented opportunity to evaluate the clinical and health economic impacts of changes in treatment patterns, potentially identifying areas in need of quality improvement programmes as well as novel insights into disease management.

## Data Availability

The aggregated results data used in this study is available for download from our website: https://wdds.ml/ with other materials such as code being accessible from repository at: https://github.com/SwanseaUniversityMedical/concept-library
The main patient-level data sources used in this study are available in the SAIL Databank at Swansea University, Swansea, UK, but as restrictions apply they are not publicly available. All proposals to use SAIL data are subject to review by an independent Information Governance Review Panel (IGRP). Before any data can be accessed, approval must be given by the IGRP. The IGRP gives careful consideration to each project to ensure proper and appropriate use of SAIL data. When access has been granted, it is gained through a privacy protecting safe haven and remote access system referred to as the SAIL Gateway. SAIL has established an application process to be followed by anyone who would like to access data via SAIL at https://www.saildatabank.com/application-process/

https://wdds.ml/

https://github.com/SwanseaUniversityMedical/concept-library

https://www.saildatabank.com/application-process/

## Declarations

### Ethics approval and consent to participate

The data used in this study are available in the SAIL Databank at Swansea University, Swansea, UK. All proposals to use SAIL data are subject to review by an independent Information Governance Review Panel (IGRP). Before any data can be accessed, approval must be given by the IGRP. The IGRP gives careful consideration to each project to ensure proper and appropriate use of SAIL data. When access has been approved, it is gained through a privacy-protecting safe haven and remote access system referred to as the SAIL Gateway. SAIL has established an application process to be followed by anyone who would like to access data via SAIL https://www.saildatabank.com/application-process. This study has been approved by SAIL Information Governance Review Panel (IGRP) under application/project number 0911 and all research conducted in this study has been completed under the permission and approval of the IGRP.

### Availability of data and materials

The aggregated results data used in this study is available for download from our website: https://wdds.ml/ with other materials such as code being accessible from repository at: https://github.com/SwanseaUniversityMedical/concept-library

The main patient-level data sources used in this study are available in the SAIL Databank at Swansea University, Swansea, UK, but as restrictions apply they are not publicly available. All proposals to use SAIL data are subject to review by an independent Information Governance Review Panel (IGRP). Before any data can be accessed, approval must be given by the IGRP. The IGRP gives careful consideration to each project to ensure proper and appropriate use of SAIL data. When access has been granted, it is gained through a privacy protecting safe haven and remote access system referred to as the SAIL Gateway. SAIL has established an application process to be followed by anyone who would like to access data via SAIL at https://www.saildatabank.com/application-process/

### Authors’ contributions

FT and AA contributed to all of the steps of the design, implementation, analysis and writing of the manuscript from inception. AA developed the proposal and the main conceptual idea. AA and LN contributed to development of mapping algorithm. FT and AA contributed to finalisation of mapping algorithm and analysis. FT designed, implemented and deployed the online tool. FT and AA jointly developed the first draft of the manuscript. RA, MG, JH and DH conceived and designed the analysis. All authors have discussed, reviewed and contributed to the final manuscript.

## Acknowledgements

This work uses data provided by patients and collected by the NHS as part of their care and support. We would also like to acknowledge all data providers who make anonymised data available for research.

We wish to acknowledge the collaborative partnership that enabled acquisition and access to the de-identified data, which led to this output. The collaboration was led by the Swansea University Health Data Research UK team under the direction of the Welsh Government Technical Advisory Cell (TAC) and includes the following groups and organisations: the Secure Anonymised Information Linkage (SAIL) Databank, Administrative Data Research (ADR) Wales, NHS Wales Informatics Service (NWIS), Public Health Wales, NHS Shared Services Partnership and the Welsh Ambulance Service Trust (WAST). All research conducted has been completed under the permission and approval of the SAIL independent Information Governance Review Panel (IGRP) project number 0911.

## Public and patient involvement

This project is undertaken under a proposal which has been submitted to the independent Information Governance Review Panel (IGRP) that includes members of the public (IGRP Project: 0911). Two members of the public are contributing to the scientific steering group of IGRP panel. Although the need for fast track analysis of these data in response to COVID-19 redirected our main focus to the research and the nature of anonymised patient data isolates the researcher from direct contact with patients involved in the study; however, the development of our online visualisation tool was intended for a lay audience. We are also intending to work closely with SAIL consumer panel group who are facilitating patient and public engagement through providing a platform for research to be presented and reviewed by members of public.

---

**Supplementary Figure 1.a:**
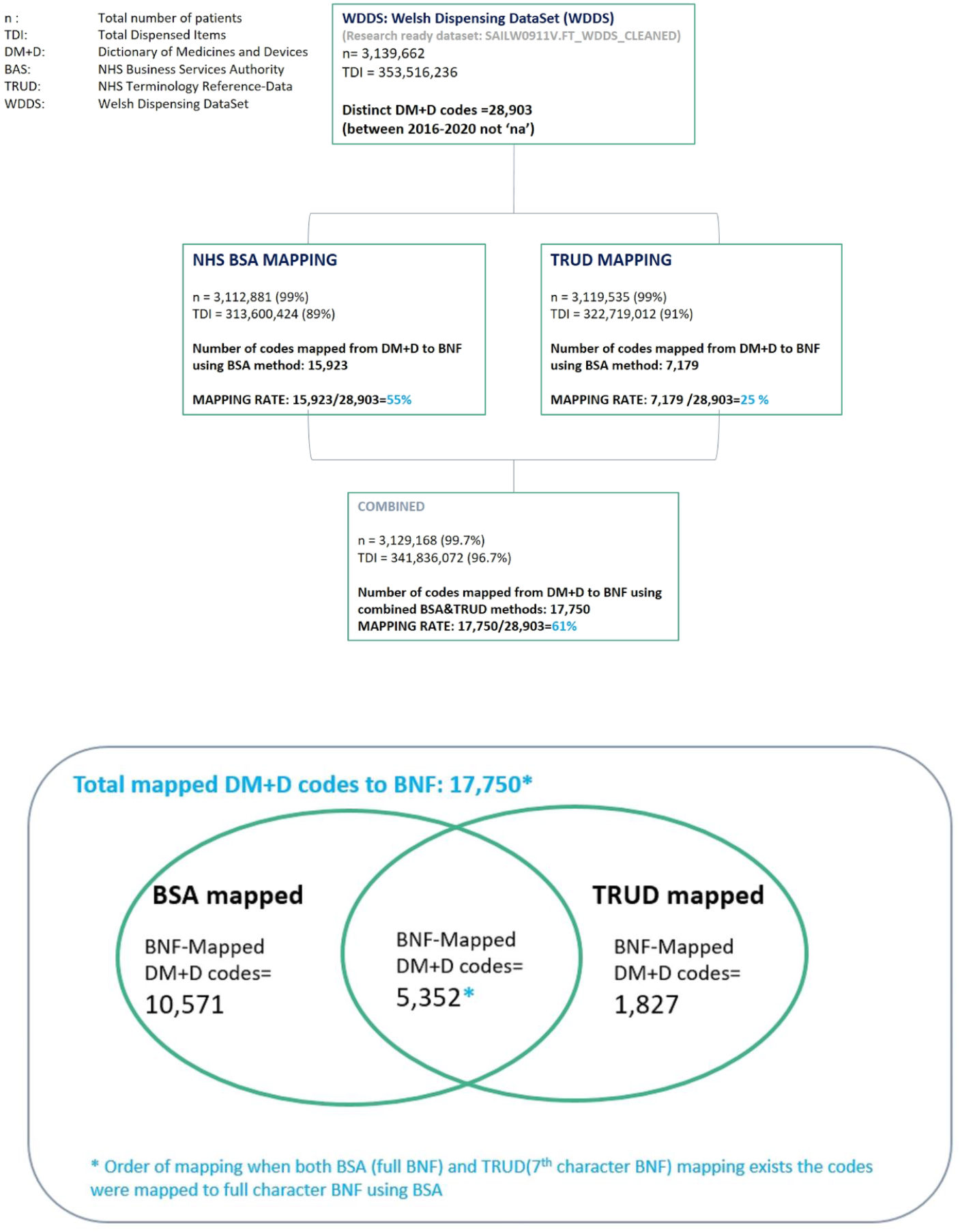
Mapping of DM+D codes from WDDS to BNF codes.

**Supplementary Figure 1.b:**
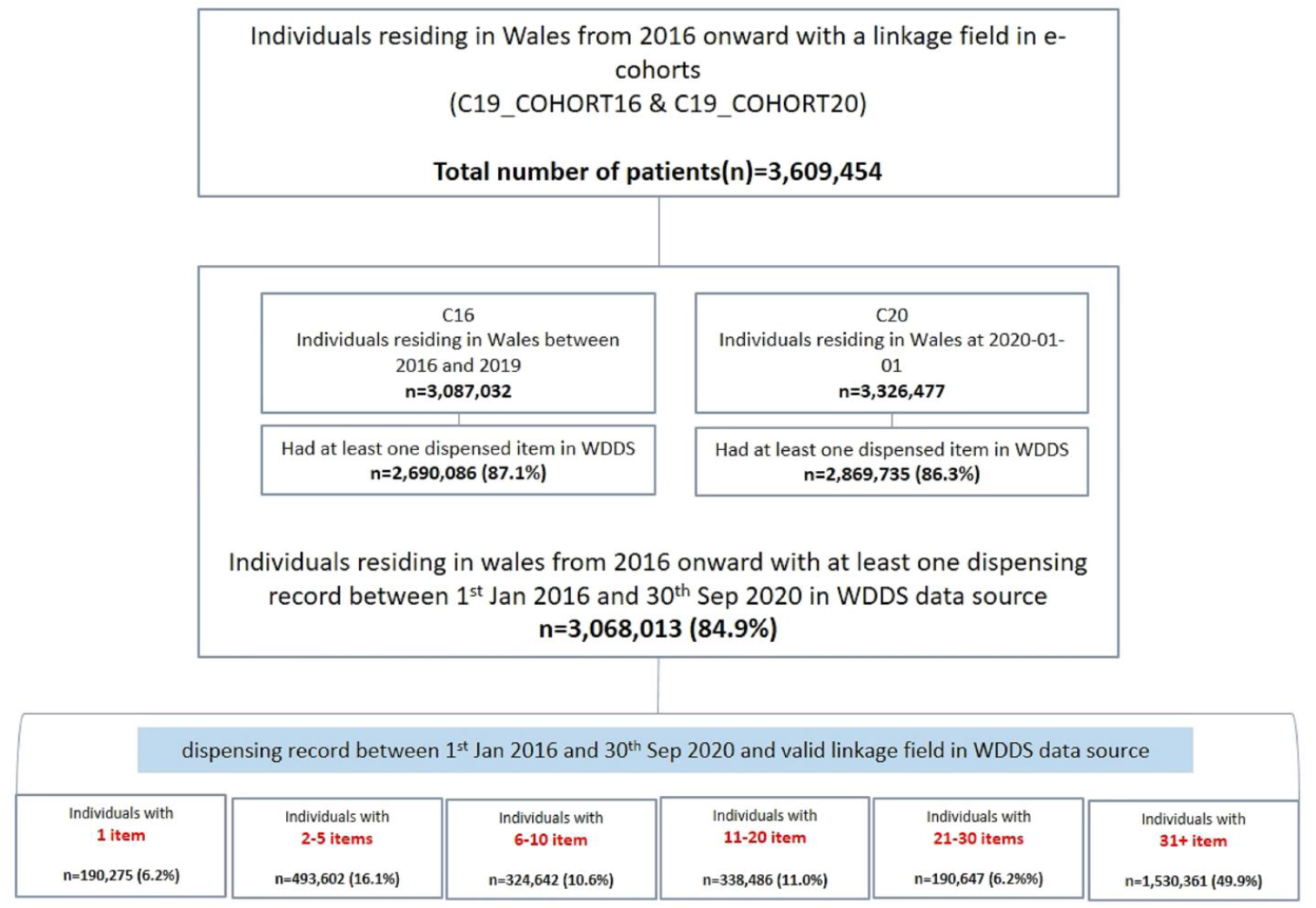
Number of dispensed items for individuals residing in Wales and linked to WDDS.

## List of abbreviations

D: Difference
RC: Relative Change
WDDS: Welsh Dispensing DataSet
C16: Electronic cohort for period of 2016-2019 (3)
C20: Electronic cohort for 2020
WDDS: Welsh Dispensing DataSet
CVS: Cardiovascular Systems
CNS: Central Nervous Systems
IVP: Immunological Products and Vaccines
NHS TRUD: Terminology Reference-Data Update Distribution
NHS BSA: NHS Business Services Authority
TDI: Total Dispensed Items
DM+D: Dictionary of medicines and devices
BNF: British National Formulary
Pop’n: Population

---

**Supplementary Figure 2.1.**
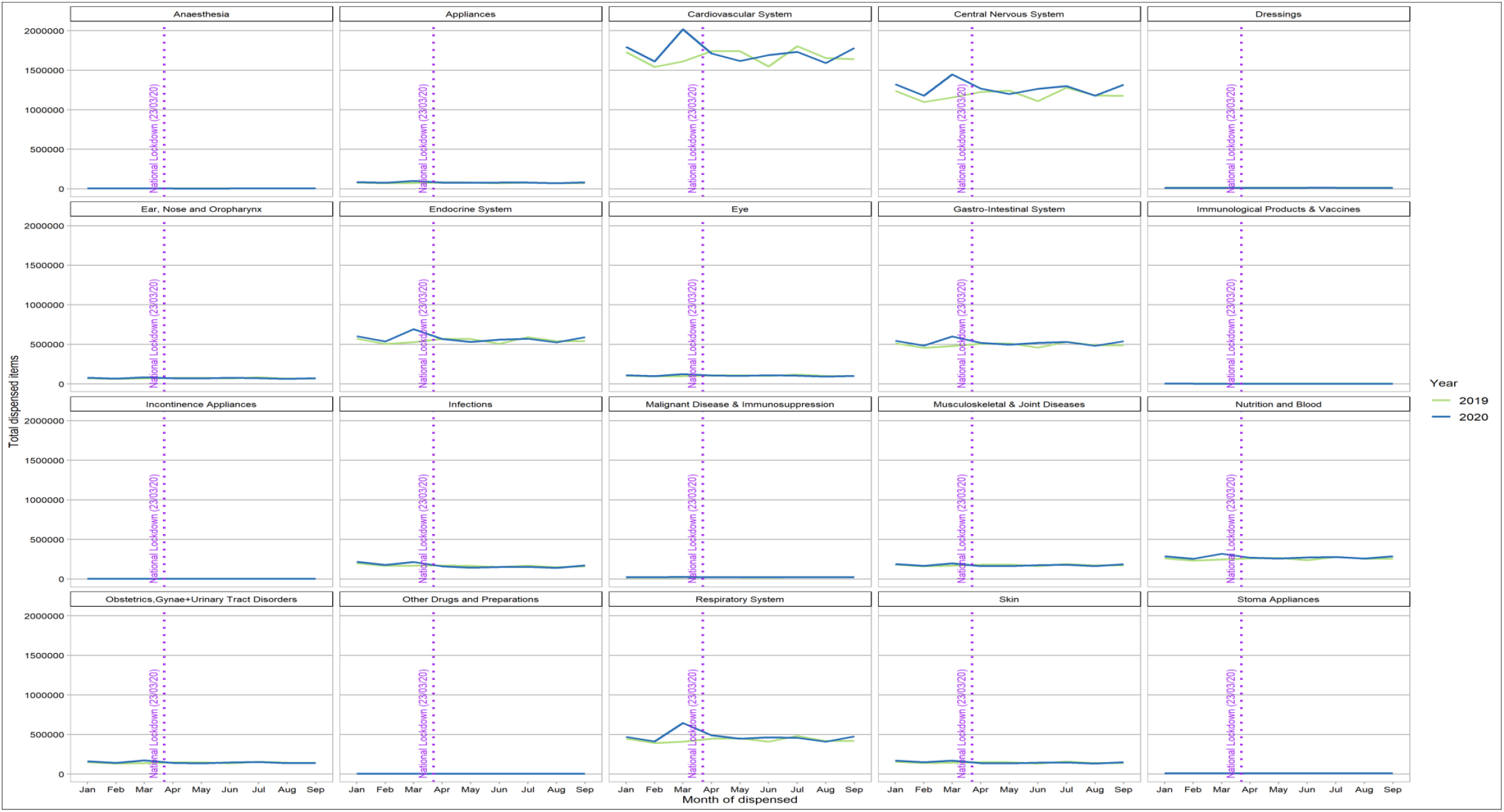
Number of dispensed items per BNF chapter-**2020 vs 2019 harmonized scale**

**Supplementary Figure 2.2.**
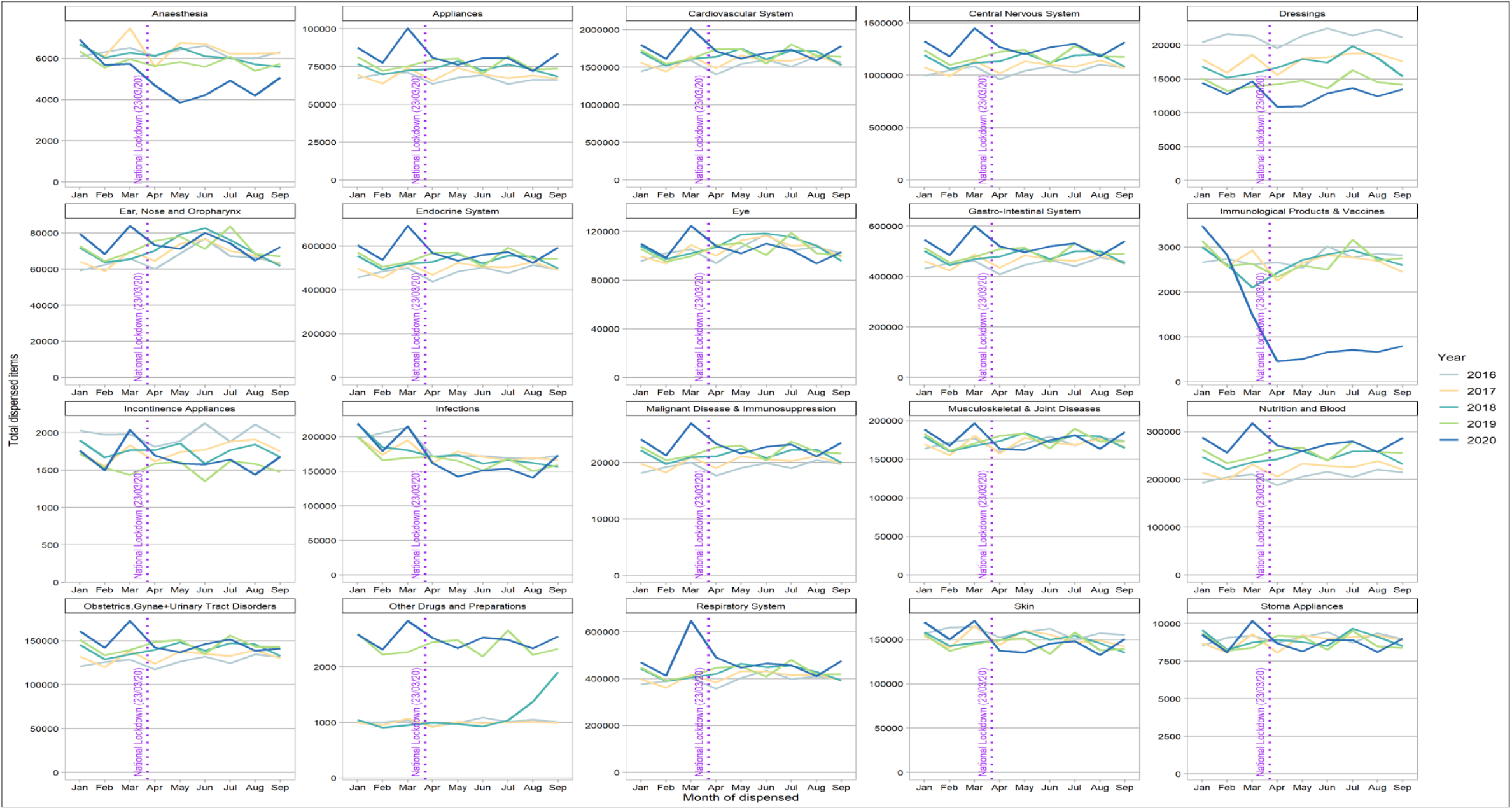
Number of dispensed items per BNF chapter per month –**in all years (numbers of each chapter are individually scaled)**

**Supplementary Figure 2.3.**
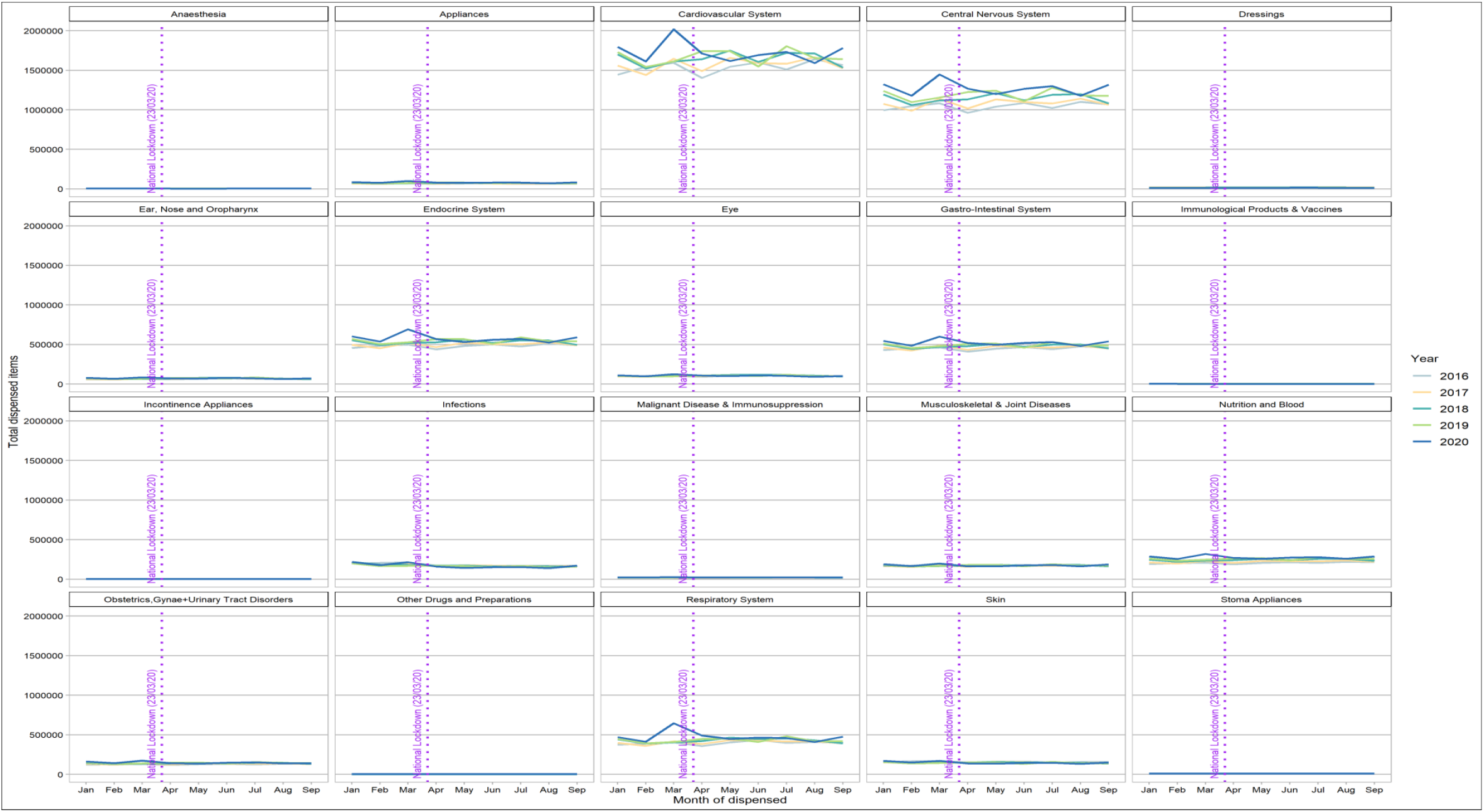
Number of dispensed items per BNF chapter per month –**in all years - harmonized scale**

**Supplementary Figure 3.1.**
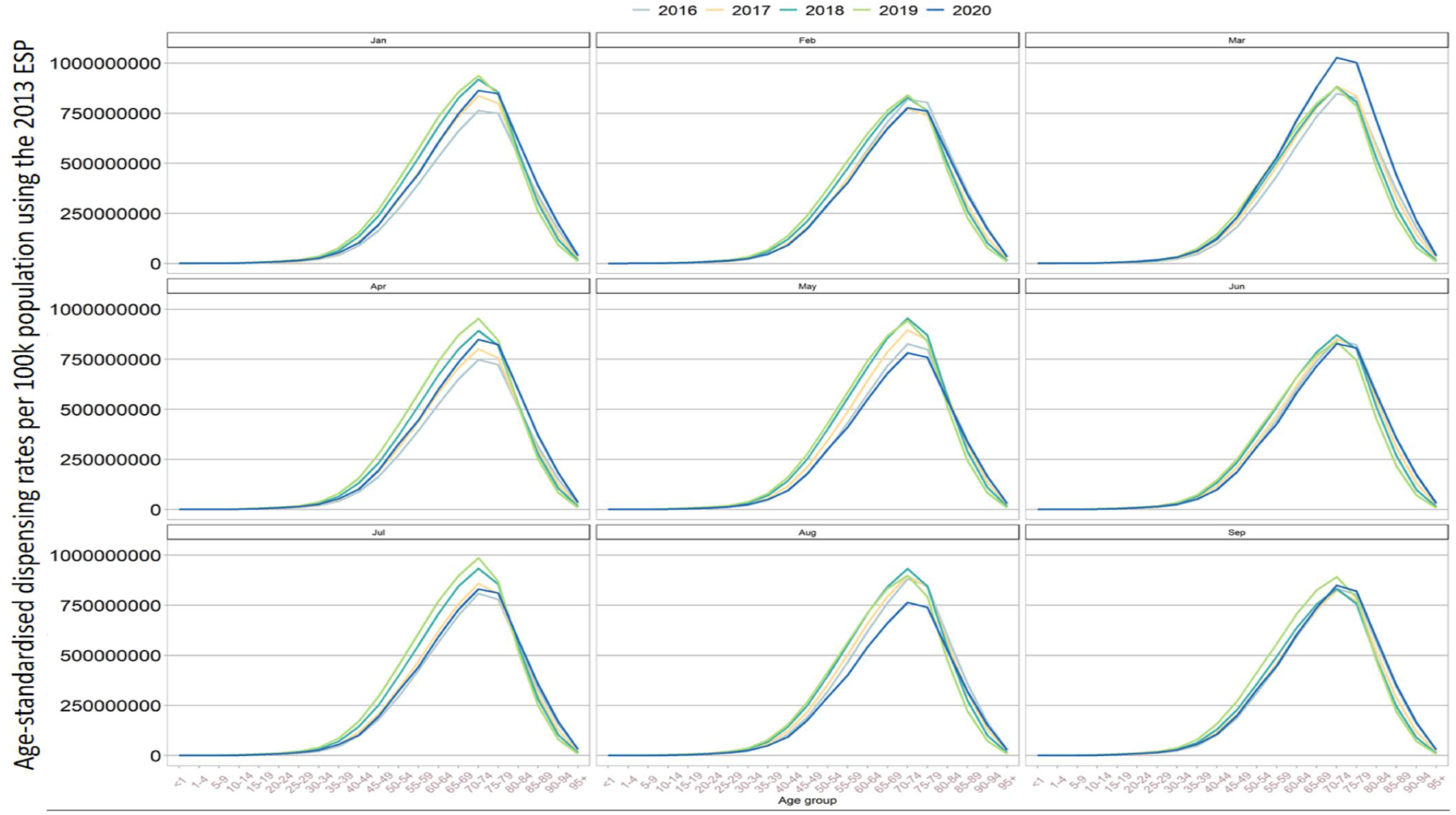
Age-standardized dispensing rates for <u>**cardiovascular system**</u> in per 100,000 pop’n per year and month

**Supplementary Figure 3.2.**
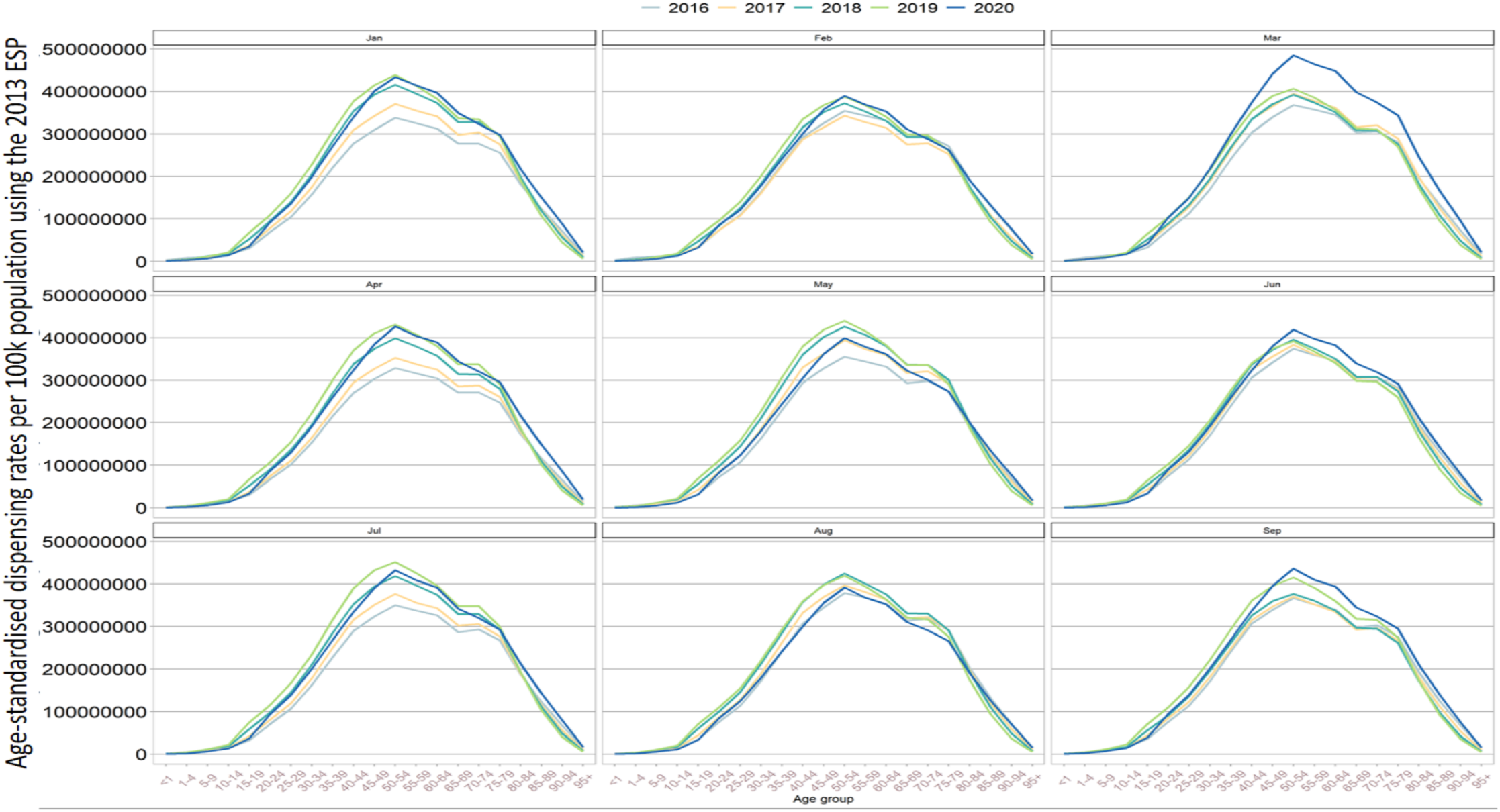
Age-standardized dispensing rates for <u>**Central Nervous System**</u> in per 100,000 pop’n per year and month

**Supplementary Figure 3.3.**
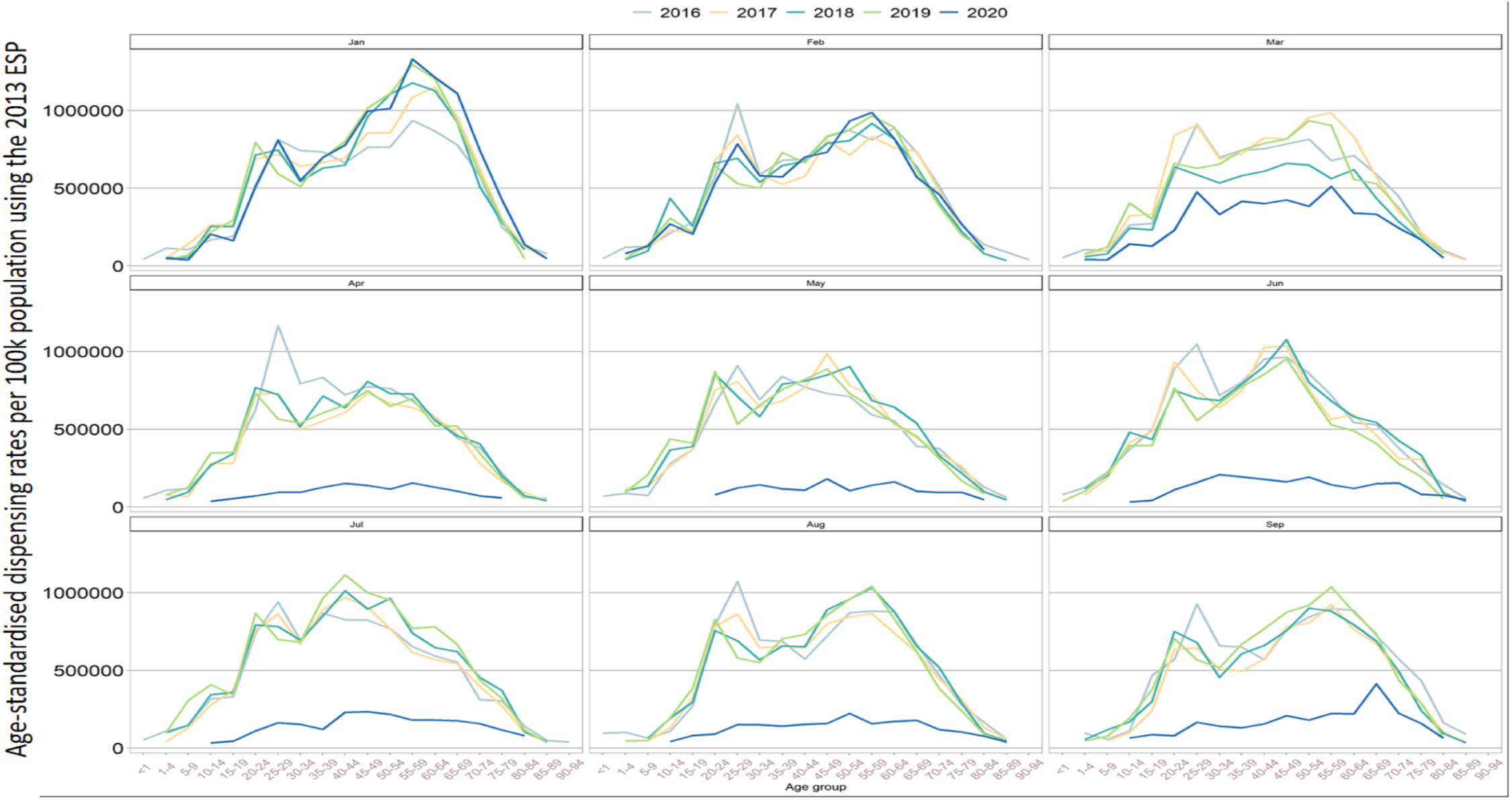
Age-standardized dispensing rates for <u>**Immunological Products & Vaccines**</u> in per 100,000 pop’n per year and month

